# Association of Systemic Immune-inflammation Index With Short-Term Mortality of Congestive Heart Failure: A Retrospective Cohort Study

**DOI:** 10.1101/2021.08.03.21261549

**Authors:** Yiyang Tang, Xiaofang Zeng, Yilu Feng, Qin Chen, Zhenghui Liu, Hui Luo, Lihuang Zha, Zaixin Yu

## Abstract

**Purpose:** The present study aimed to clarify the potential predictive significance of Systemic immune-inflammation index (SII) in assessing the poor prognosis of critically ill patients with congestive heart failure (CHF).

**Methods:** Detailed clinical data were extracted from the Multiparameter Intelligent Monitoring in Intensive Care III database after gaining access and building the local platform. The 30- and 90-day and hospital all-cause mortalities of the patient was the primary outcome, and the readmission rate and the occurrence of major cardiovascular adverse events (MACEs) were the secondary outcomes. the Cox proportional hazard model and Logistic regression analysis were selected to reveal the relationship between SII level and the research outcome. Further, the propensity score matching (PSM) analysis was performed to improve the reliability of results by reducing the imbalance across groups.

**Results:** There were a total of 4606 subjects who passed the screening process and entered the subsequent analysis. Multivariate regression analysis showed that after adjusting for possible confounders, including age, heart rate, and albumin, etc., the high level of SII was independently associated with 30- and 90-day and hospital mortalities (tertile 3 versus tertile 1: HR, 95% CIs: 1.23, 1.04-1.45; 1.21, 1.06-1.39; 1.26, 1.05-1.50) and the incidence of MACEs (tertile 3 versus tertile 1: OR, 95% CI: 1.39, 1.12-1.73) in critically ill patients with CHF, but no significant correlation was found between SII and the readmission rate. Consistently, patients with high SII level still presented a significantly higher short-term mortality than patients with low SII in the PSM subset.

**Conclusion:** In critically ill patients with CHF, high level of SII could effectively predict high 30- and 90-day and hospital mortalities, as well as the high risk of occurrence of MACEs.

## 1 Introduction

Congestive heart failure (CHF) is defined as a pathological condition in which the cardiac output is insufficient to maintain the perfusion and metabolic needs of various tissues and organs, mainly characterized by the congestion of pulmonary circulation, accompanied by the corresponding clinical manifestations such as shortness of breath and decreased activity tolerance(Sun et al., 2020). CHF is the severe or terminal stage of most primary cardiovascular diseases and has increasingly become a major public problem threatening human health. In the United States and Europe, more than 1 million heart failure patients are hospitalized each year, and as the population ages, this number is expected to increase by more than 50% in the next 15 years(Povsic, 2018). In addition, although some progress has been made in the treatment in recent years, the prognosis of CHF patients is still poor, with 5-year mortality rate as high as 40-50%(Chen et al., 2011). Therefore, accurate assessment and stratification of prognosis are critical to the clinical management of CHF, and the development of certain prognostic-related biomarkers is also pressing, which can help clinicians to identify high-risk patients early and take more aggressive treatment measures(Xu et al., 2021).

Although the specific pathogenesis of heart failure remains unclear, abnormal immune activation and chronic inflammation play an important role. The damage, repair and remodeling of myocardium are the important link in the occurrence and development of heart failure, of which immune/inflammatory cells (neutrophils, lymphocytes, etc.) and the inflammatory factors (tumor necrosis factor, interleukin-6, etc.) released by them are involved(Li et al., 2021). Inflammatory factors can induce cardiomyocyte hypertrophy, apoptosis and fibrosis, and ultimately lead to adverse cardiac remodeling and progressive left ventricular dysfunction(Bradham et al., 2002). Inflammatory factors have been considered to be biomarkers for poor prognosis of heart failure, and anti-inflammatory therapy is also expected to become a new target for the treatment of heart failure(Mann, 2015).

Systemic immune-inflammation index (SII) is a composite inflammatory indicator that combines three significant immune cells, including neutrophil, lymphocyte, and platelet, and is considered to be an excellent indicator of local immune response and systemic inflammation(Wang et al., 2016). Neutrophils, platelets and the cytokines they produce are mainly related to non-specific immune responses, while lymphocytes are considered to be mainly related to specific immune pathways. Compared with the absolute count of single immune cells, SII has a better representation in reflecting the inflammation status of the body, with better stability. Until now, SII has been confirmed to be closely correlated with the poor prognosis of a variety of cardiovascular diseases, including coronary artery disease(Yang et al., 2020), aortic stenosis(Tosu et al., 2021), infective endocarditis(Agus et al., 2020), etc., showing good application prospects, but no studies have shown that SII is correlated with the prognosis of patients with CHF. In view of this, the present study attempted to elucidate the independent association between SII and the occurrence of adverse events such as short-term mortality in critically ill patients with CHF, so as to provide reference for the prevention and treatment of heart failure.

## 2 Materials and Methods

### 2.1 Data Sources

All data used in the research and analysis of this study are derived from MIMIC-Ⅲ (Medical Information Mart for Intensive Care Ⅲ) database, a free and open-access large-scale critical care medicine database developed and run by the Massachusetts Institute of Technology for researchers all over the world. Another advantage of this database, in addition to including the detailed clinical data such as demographic data, vital signs, laboratory tests, and various treatment information, is that it provides accurate death information of all subjects, including time of death in the hospital or within 90 days after discharge, which makes it possible for the clinicians to carry out the prognostic related research. Besides, since this database has been approved by the local ethics committee as a whole and the individual identifying information of the research subjects has been deleted, this study no longer needs additional ethical approval.

### 2.2 Population Selection and Exclusion

The subjects of this study are all from the MIMIC-Ⅲ database(Johnson et al., 2016), and patients who meet all the following requirements are included in the subsequent analysis: (1) Patients diagnosed with CHF based on the ICD-9 code (code 428.0); (2) Adult patients 18 years of age and older; (3) First admission to ICU. Patients who met one of the following criteria were excluded from the study: (1) The length of ICU stay was shorter than 24 hours; (2) Absence of SII results during hospitalization; (3) Survival time was less than 0 (the time of death of some organ donors may be earlier than the admission).

### 2.3 Study Outcomes

The short-term mortality, including the 30- and 90-day and hospital all-cause mortalities of critically ill patients with CHF, were selected as the primary study outcome, while the secondary study outcome events were defined as the patients’ readmission rate and the occurrence of major adverse cardiac events (MACEs), which is a composite outcome event, including all-cause death, readmission for acute heart failure, the use of mechanical circulatory support and the implementation of heart transplantation(Li et al., 2020).

### 2.4 Data Extraction and Integration

Through the PostgreSQL software (version 9.6, https://www.postgresql.org/), we extracted detailed clinical data of the research subjects from the MIMIC-Ⅲ database, including demographic data (age, gender, race, etc.), comorbidities (hypertension, diabetes mellitus, atrial fibrillation, etc.), vital signs (heart rate, blood pressure, respiratory rate, etc.), severity of illness scores (SOFA and SAPSII scores), laboratory tests (blood routine, electrolytes, etc.) and intervention measures (dialysis, mechanical ventilation, etc.). And the value of SII is equal to the product of the platelet count and the neutrophil count divided by the lymphocyte count(Hu et al., 2014).

After separately extracting the required tables from the MIMIC-Ⅲ database, the Stata software (version 16, https://www.stata.com/) was used to process and merge these original table and generate a complete table that can be used for subsequent analysis. Use the “winsorize” function to identify and process outliers, and fill in missing values through multiple compensation methods.

### 2.5 Statistical Analyses

The continuous variables were presented in the form of mean ± standard deviation (SD) or median (interquartile range). If continuous variables satisfy both normal distribution and homogeneity of variance, t-test was used for analysis, while the Mann-Whitney U test was conducted if the normal distribution or homogeneity of variance were not satisfied. Categorical variables are expressed in the form of the number of cases (percentage), with the chi-square test (or Fisher’s exact method) for analysis.

We constructed a generalized additive model (GAM) to determine the non-linear relationship between SII and 30- and 90-day all-cause mortalities in critically ill patients with CHF. In addition, we also visually show the relationship between SII and patients’ survival through the Kaplan-Meier (K-M) curve, and use the Log-rank test for hypothesis testing.

In order to relieve the interference of possible confounding factors on the results, we completed univariate and multivariate regression analysis to further clarify the relationship between SII and outcome variables. In the crude model, no variables were adjusted. In model I, the age, gender, and race variables were adjusted, while model II further adjusted other 13 variables on the basis of these variables of model II, including heart rate, blood urea nitrogen (BUN), albumin, troponin T (cTnT), N-terminal probrain natriuretic peptide (NT-proBNP), urine output of first day, cardiac index, the type of first ICU admission, the use of mechanical ventilation and vasopressor, pneumonia, and liver diseases. The selection of confounding factors follows the following principles(Jaddoe et al., 2014): (1) A certain factor has an influence of more than 10% on the research variable; (2) Some factors may have a significant impact on the outcome variable based on past experience. Besides, we used the variance inflation factor (VIF) to test the multicollinearity between variables with 5 as the threshold, and the variables with a high degree of collinearity, serum chloride, were deleted to avoid over-fitting of the model. The Cox regression analysis was used to determine the relationship between SII and short-term mortalities in critically ill patients with CHF, while Logistic regression analysis was used to analyze the association between SII and readmission rates or MACEs.

To further enhance the credibility of our analysis, we performed the subgroup analysis and propensity score matching (PSM) analysis. First, By the subgroup analysis, determine whether the correlation between the SII and the high 30-day all-cause mortality in critically ill patients with CHF was consistent across various subgroups stratified mainly by comorbidities, and reflect the stability of SII as a prognostic marker. The optimal cut-off value of SII for the short-term mortality was determined by the X-tile (version 3.6.1, Yale University School of Medicine) software, and then the entire study population was divided into two groups, namely the high SII group and the low SII group. In this study, all variables with uneven distribution between the two groups of patients were included in the PSM model as covariates, and the corresponding propensity score was calculated by Logistic regression. Then the two groups of individuals with similar propensity scores were matched by 1:1 using nearest neighbor matching method with a caliper width of 0.05, and the histogram of the propensity score distribution was draw, which was carried out through the Stata software. Then, K-M curves were depicted and Cox regression analysis was conducted in the matching cohort after PSM, which further validated SII as an independent risk factor for poor prognosis in the critically ill patients with CHF.

The statistical analysis above was conducted by EmpowerStats software (version 2.20, http://www.empowerstats.com/cn/, X&Y solutions, Inc, Boston, MA) and R software (version 3.4.3). p <0.05 (two-sided) was considered statistically significant.

## 3 Results

### 3.1 Clinical Characteristics of Study Subjects

The study subjects were screened according to the process mentioned above, and 4606 subjects were finally included. The demographic data, vital signs, comorbidities, treatment, scores and laboratory examinations and other relevant information are shown in Table 1 in detail. In general, the study subjects included were a little old, with a median age of 74.91 years old, mainly male patients, reaching 2436, accounting for 52.89%. And the majority of the subjects were whites, accounting for 72.71%, blacks only 7.21%, and other races including Asians accounted for 20.08%. The duration of follow-up was 90 days, of which 3226 survived and 1380 died due to various reasons before the end of the study. SII (213.97 vs 163.11, p<0.001) was significantly higher in the non-survivor cohort. Compared with the survivor group, older patients were identified more frequently in the non-survivor group, with more unstable vital signs, higher incidence of comorbidities such as VHD, atrial fibrillation, and renal failure, as well as higher SOFA and SAPSII scores.

**Table 1.**
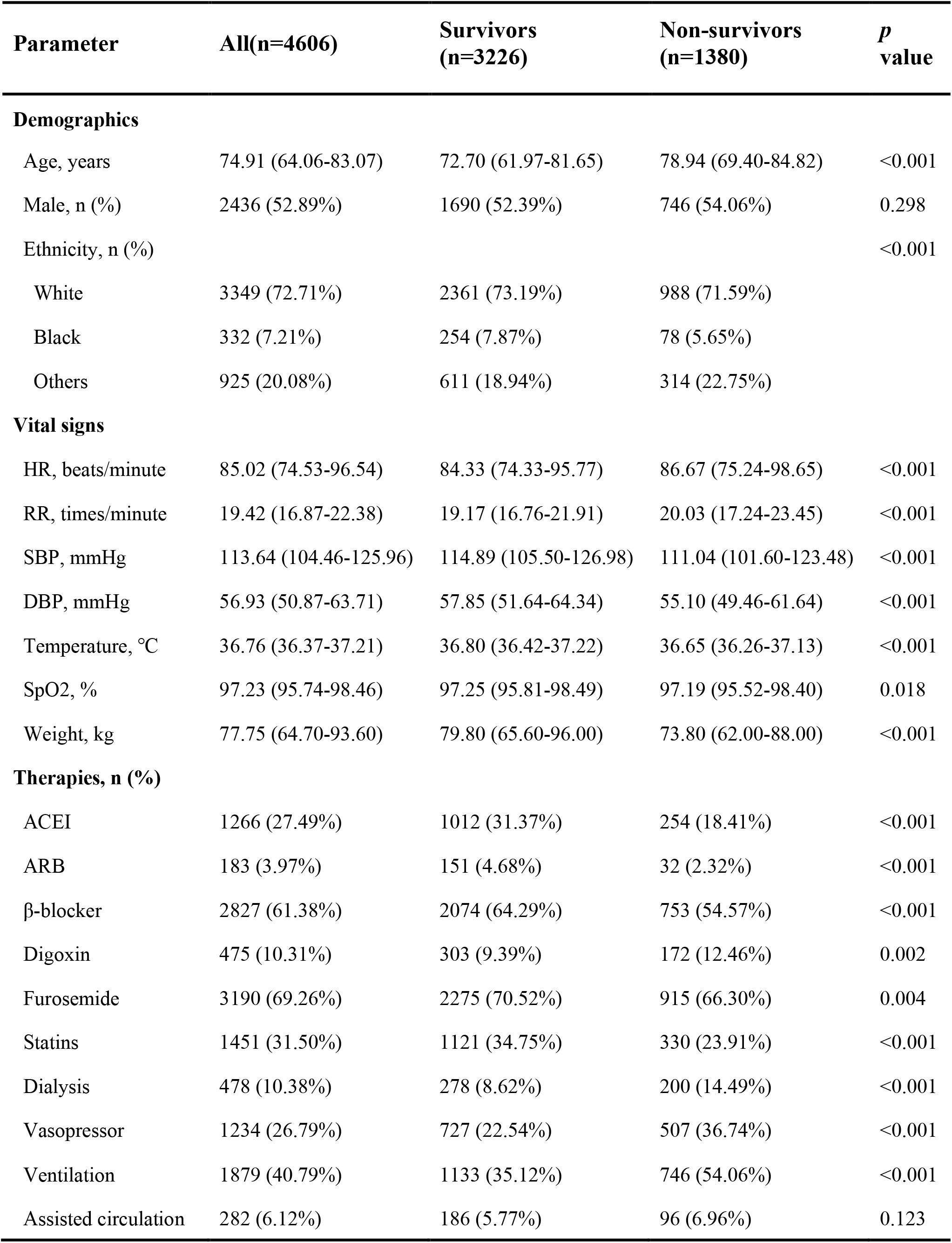
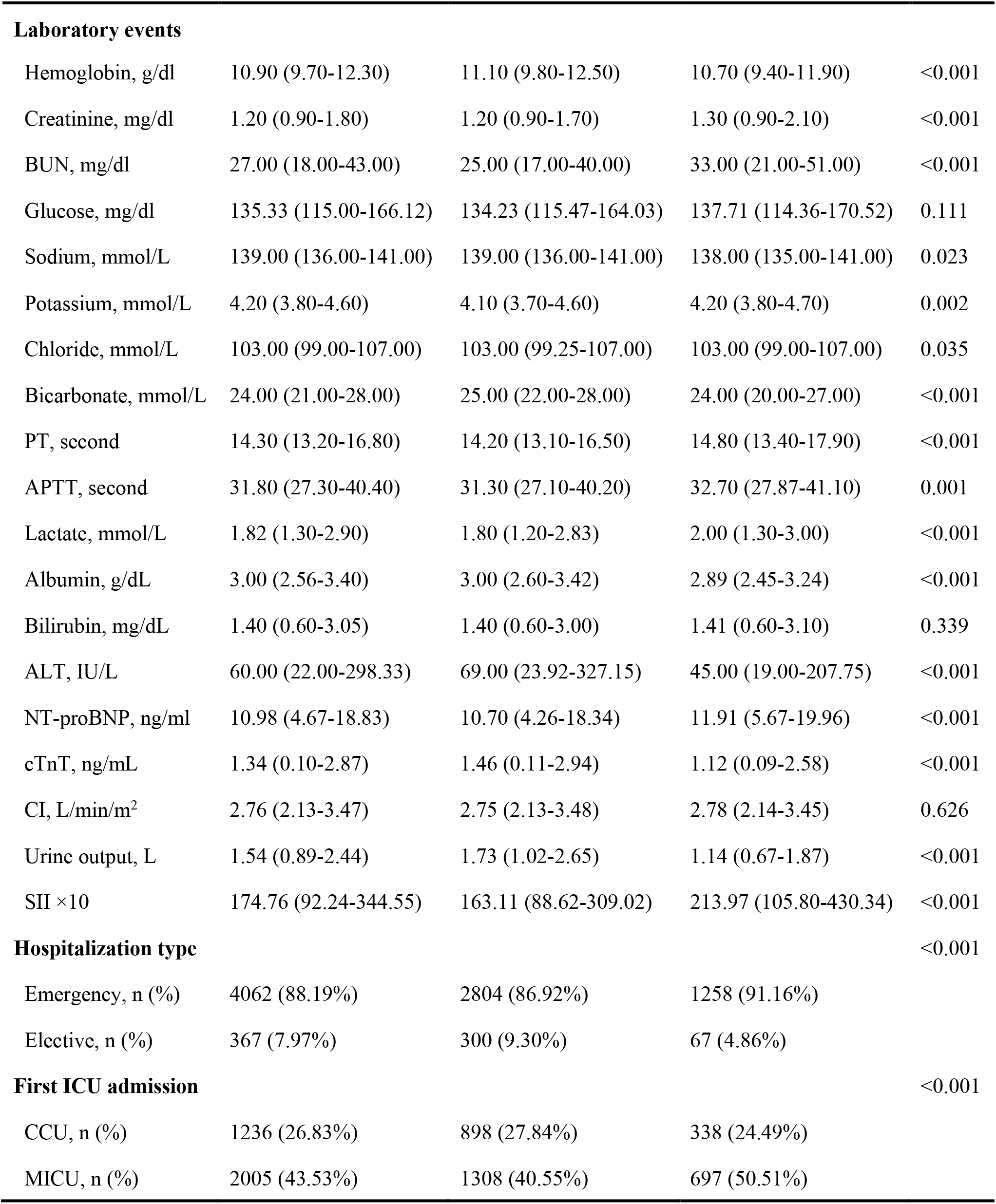
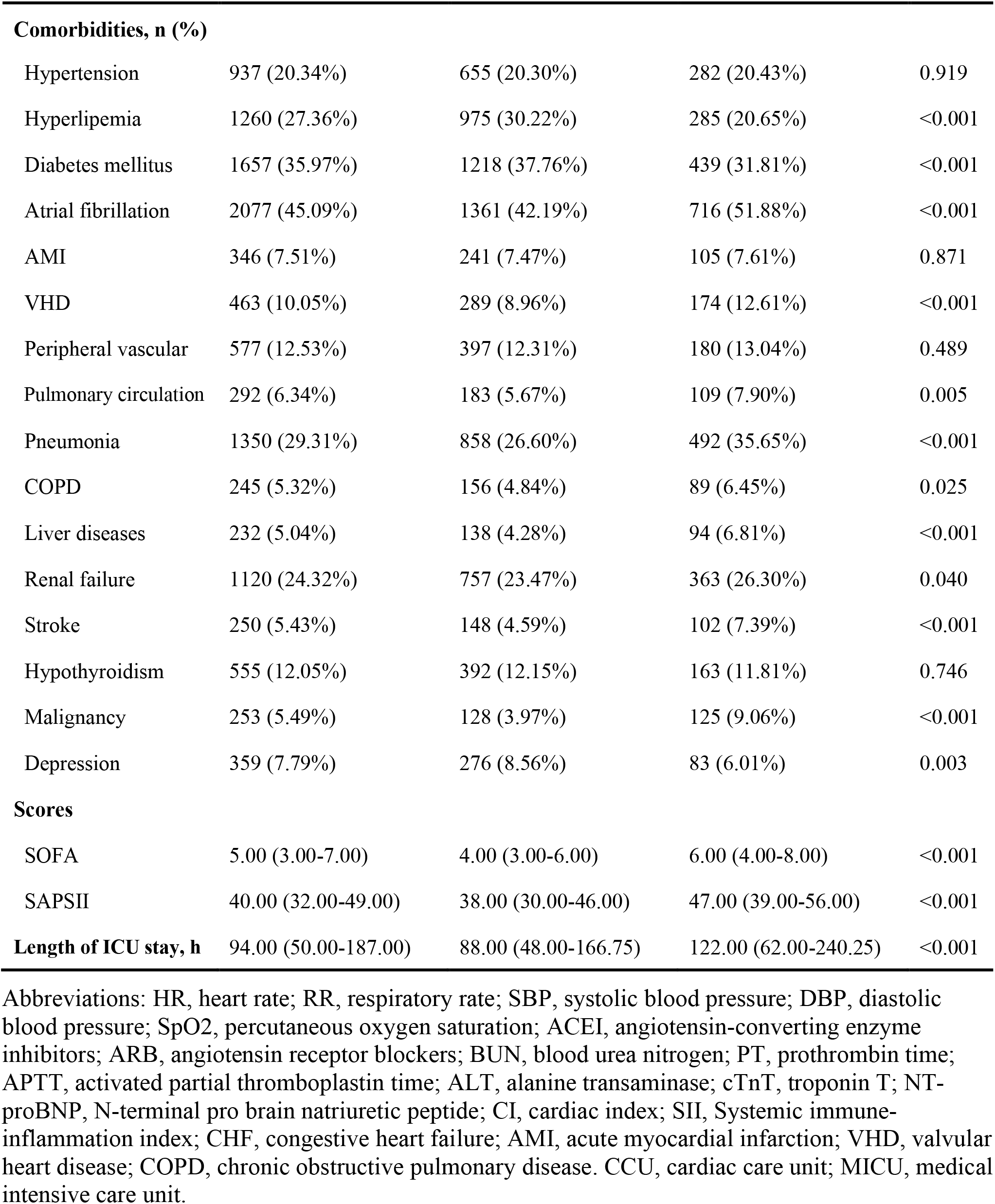
The baseline clinical characteristics of critically ill patients with CHF.

### 3.2 SII Distinguishing a Poor Short-term Prognosis

GAM analysis was used to identify the non-linear correlation between SII and the short-term all-cause mortality of patients, which showed that there was a U-shaped curve relationship between SII and 30-day (Figure 1A) and 90-day (Figure 1B) all-cause mortalities in critically ill patients with CHF, indicating that higher or lower SII may be associated with increased mortality. The K-M survival curve (Figure 2) also showed that compared with the low SII group, patients with high SII level had a lower overall survival rate and shorter survival time, with a statistically significant *p* value.

**Figure 1.**
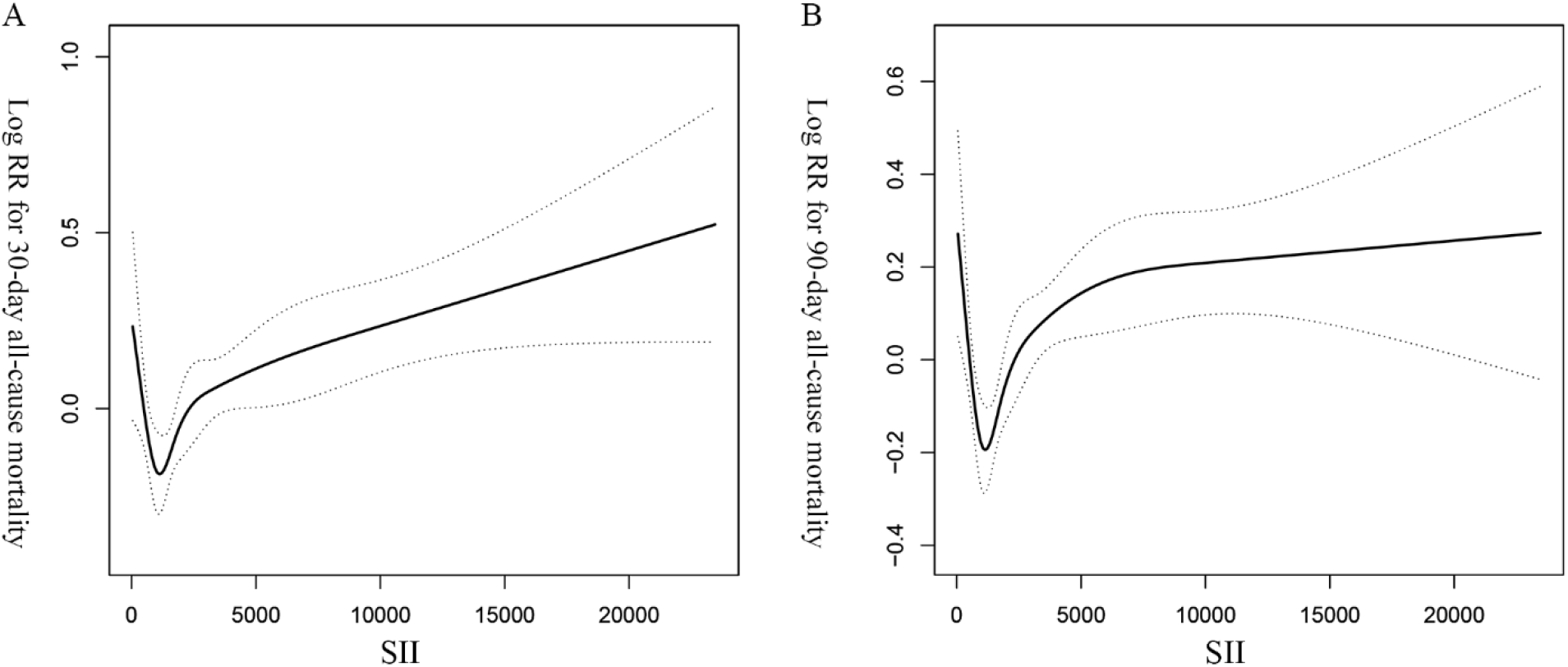
The non-linear curve fitting of the association between SII and 30- (A) and 90- (B) day all-cause mortalities in critically ill patients with CHF after adjusting for age, gender, race, and other potential variables. SII, systemic immune-inflammation index. CHF, congestive heart failure.

**Figure 2.**
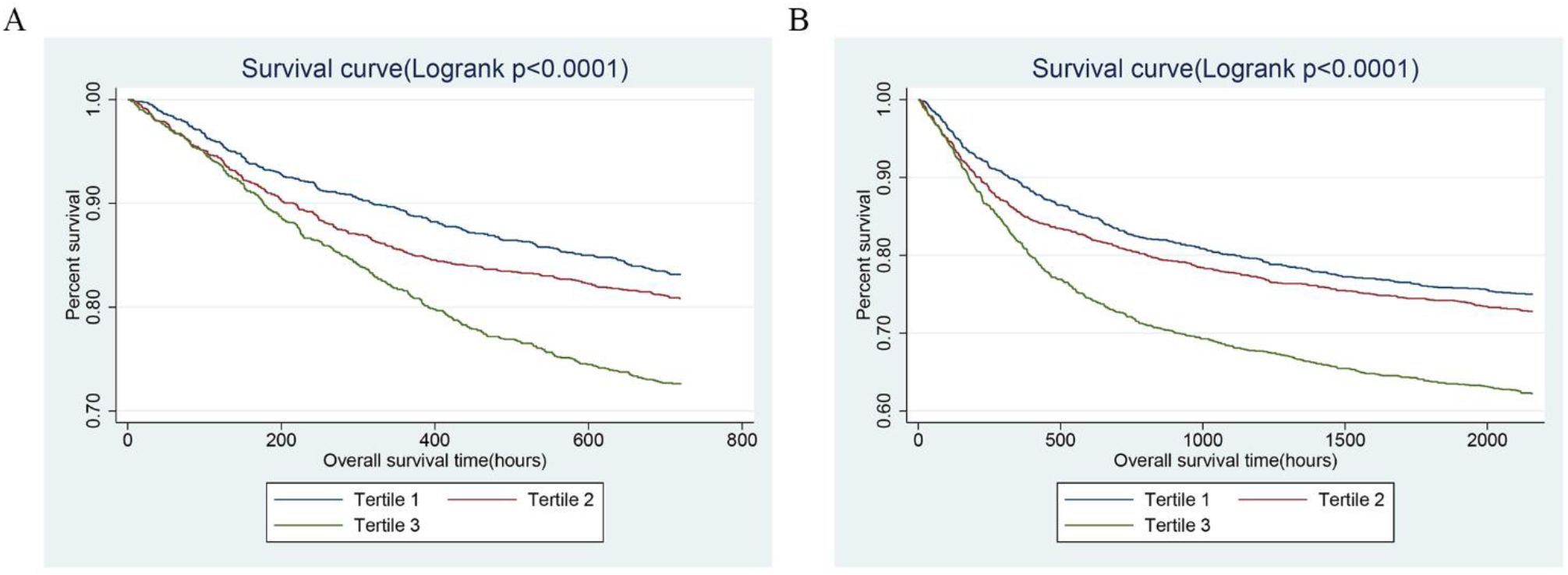
Kaplan-Meier survival curves showing the 30- (A) and 90- (B) day all-cause mortalities stratified by SII tertile in critically ill patients with CHF. SII, systemic immune-inflammation index. CHF, congestive heart failure.

In order to verify the independent relationship between SII and the poor short-term mortality of CHF patients, we performed the univariate and multivariate Cox regression analysis (Table 2), with SII stratified by tertiles. In crude model, the third tertile of SII increased significantly the risk of 30- (HR, 95% CI: 1.73, 1.48-2.03) and 90-day (HR, 95% CI: 1.65, 1.45-1.88) and hospital (HR, 95% CI: 1.61, 1.36-1.91) all-cause mortalities compared with the first tertile. In multivariate model I, after adjusting for age, sex, and race, the third tertile of SII group also suffered from the higher risk of 30- (HR, 95% CI: 1.63, 1.40-1.91) and 90-day (HR, 95% CI: 1.57, 1.38-1.79) and hospital (HR, 95% CI: 1.45, 1.22-1.72) all-cause mortalities. In model II, in addition to adjusting for the variables in model I and other possible confounders such as pneumonia, the use of dialysis, and BUN level, high levels of SII were still strongly associated with poor 30- (HR, 95% CI: 1.23, 1.04-1.45) and 90-day (HR, 95% CI: 1.21, 1.06-1.39) and hospital (HR, 95% CI: 1.26, 1.05-1.50) all-cause mortalities.

**Table 2.** The univariate and multivariate Cox regression analysis exploring the association of SII tertile with short-term mortality of critically ill patients with CHF.

|  | Crude |  | Model I |  | Model II |  |
| --- | --- | --- | --- | --- | --- | --- |
|  | HR (95% CIs) | P value | HR (95% CIs) | P value | HR (95% CIs) | P value |
| 30-day all-cause mortality |  |  |  |  |  |  |
| SII | 1.00 (1.00, 1.00) | <0.001 | 1.00 (1.00, 1.00) | <0.0001 | 1.00 (1.00, 1.00) | <0.0001 |
| SII (tertile) |  |  |  |  |  |  |
| <1144.28 | 1 (ref) |  | 1 (ref) |  | 1 (ref) |  |
| ≥1144.28, <2730.11 | 1.17 (0.99, 1.38) | 0.067 | 1.14 (0.97, 1.35) | 0.112 | 1.05 (0.89, 1.25) | 0.524 |
| ≥2730.11 | 1.73 (1.48, 2.03) | <0.001 | 1.63 (1.40, 1.91) | <0.001 | 1.23 (1.04, 1.45) | 0.011 |
| P for trend | <0.001 |  | <0.001 |  | <0.001 |  |
| 90-day all-cause mortality |  |  |  |  |  |  |
| SII | 1.00 (1.00, 1.00) | <0.001 | 1.00 (1.00, 1.00) | <0.0001 | 1.00 (1.00, 1.00) | 0.001 |
| SII (tertile) |  |  |  |  |  |  |
| <1144.28 | 1 (ref) |  | 1 (ref) |  | 1 (ref) |  |
| ≥1144.28, <2730.11 | 1.12 (0.97, 1.28) | 0.114 | 1.10 (0.96, 1.27) | 0.168 | 1.02 (0.88, 1.17) | 0.820 |
| ≥2730.11 | 1.65 (1.45, 1.88) | <0.001 | 1.57 (1.38, 1.79) | <0.001 | 1.21 (1.06, 1.39) | 0.005 |
| P for trend | <0.001 |  | <0.001 |  | <0.001 |  |
| Hospital all-cause mortality |  |  |  |  |  |  |
| SII | 1.00 (1.00, 1.00) | <0.001 | 1.00 (1.00, 1.00) | <0.0001 | 1.00 (1.00, 1.00) | 0.002 |
| SII (tertile) |  |  |  |  |  |  |
| <1144.28 | 1 (ref) |  | 1 (ref) |  | 1 (ref) |  |
| ≥1144.28, <2730.11 | 1.27 (1.06, 1.52) | 0.008 | 1.21 (1.01, 1.45) | 0.035 | 1.17 (0.97, 1.40) | 0.096 |
| ≥2730.11 | 1.61 (1.36, 1.91) | <0.001 | 1.45 (1.22, 1.72) | <0.001 | 1.26 (1.05, 1.50) | 0.011 |
| P for trend | <0.001 |  | <0.001 |  | <0.001 |  |

Crude model was adjusted for none.
Model I was adjusted for age, gender, and race.
Model II was adjusted for age, gender, race, heart rate, blood urea nitrogen, albumin, troponin T, NT-proBNP, urine output of first day, cardiac index, the type of first ICU admission, the use of mechanical ventilation and vasopressor, pneumonia, and liver diseases.
Abbreviations: SII, Systemic immune-inflammation index; CHF, congestive heart failure; HR, hazard ratio; CI, confidence interval.

### 3.3 Association between SII and Readmission and MACEs

For MACEs, a similar trend was observed. In model II (Table 3), patients with an SII ≥2730.11 were still at higher incidence of MACEs (OR, 95% CI: 1.39, 1.12-1.73). However, in the multivariate regression analysis, there was no obvious independent correlation between SII and readmission rate in the critically ill patients with CHF (tertile 3 versus tertile 1: OR, 95% CI: 0.97, 0.81-1.15).

**Table 3.** The univariate and multivariate Logistic regression analysis exploring the association of SII tertile with readmission and MACEs of critically ill patients with CHF.

|  | Crude |  | Model I |  | Model II |  |
| --- | --- | --- | --- | --- | --- | --- |
|  | OR (95% CIs) | P value | OR (95% CIs) | P value | OR (95% CIs) | P value |
| Readmission |  |  |  |  |  |  |
| SII (tertile) |  |  |  |  |  |  |
| <1144.28 | 1 (ref) |  | 1 (ref) |  | 1 (ref) |  |
| ≥1144.28, <2730.11 | 0.91 (0.77, 1.07) | 0.253 | 0.94 (0.80, 1.11) | 0.472 | 0.94 (0.80, 1.11) | 0.484 |
| ≥2730.11 | 0.88 (0.75, 1.04) | 0.129 | 0.93 (0.78, 1.10) | 0.392 | 0.97 (0.81, 1.15) | 0.703 |
| P for trend | <0.001 |  | <0.001 |  | <0.001 |  |
| MACEs |  |  |  |  |  |  |
| SII (tertile) |  |  |  |  |  |  |
| <1144.28 | 1 (ref) |  | 1 (ref) |  | 1 (ref) |  |
| ≥1144.28, <2730.11 | 1.17 (1.02, 1.35) | 0.030 | 1.17 (1.02, 1.35) | 0.028 | 0.92 (0.74, 1.13) | 0.417 |
| ≥2730.11 | 2.01 (1.73, 2.32) | <0.001 | 2.02 (1.74, 2.35) | <0.001 | 1.39 (1.12, 1.73) | 0.003 |
| P for trend | <0.001 |  | <0.001 |  | <0.001 |  |

Crude model was adjusted for none.
Model I was adjusted for age, gender, and race.
Model II was adjusted for age, gender, race, heart rate, blood urea nitrogen, albumin, troponin T, NT-proBNP, urine output of first day, cardiac index, the type of first ICU admission, the use of mechanical ventilation and vasopressor, pneumonia, and liver diseases.
Abbreviations: SII, Systemic immune-inflammation index; CHF, congestive heart failure; HR, hazard ratio; CI, confidence interval.

### 3.4 Subgroup Analyses

The subgroup analysis was conducted to reveal the correlation between SII and 30-day mortality across comorbidities and different parameters, and the results were shown in Table 4. First, the results of the study showed that in all subgroups, the increase in SII level was closely related to the increase in the 30-day all-cause mortality of critically ill patients with CHF. Besides, most of the stratification factors have not been found to have a significant impact on the relationship between the SII and 30-day all-cause mortality (interaction p value>0.05), except for hyperlipemia (*p*=0.006) and acute myocardial infarction (AMI, *p*= 0.016).

**Table 4.**
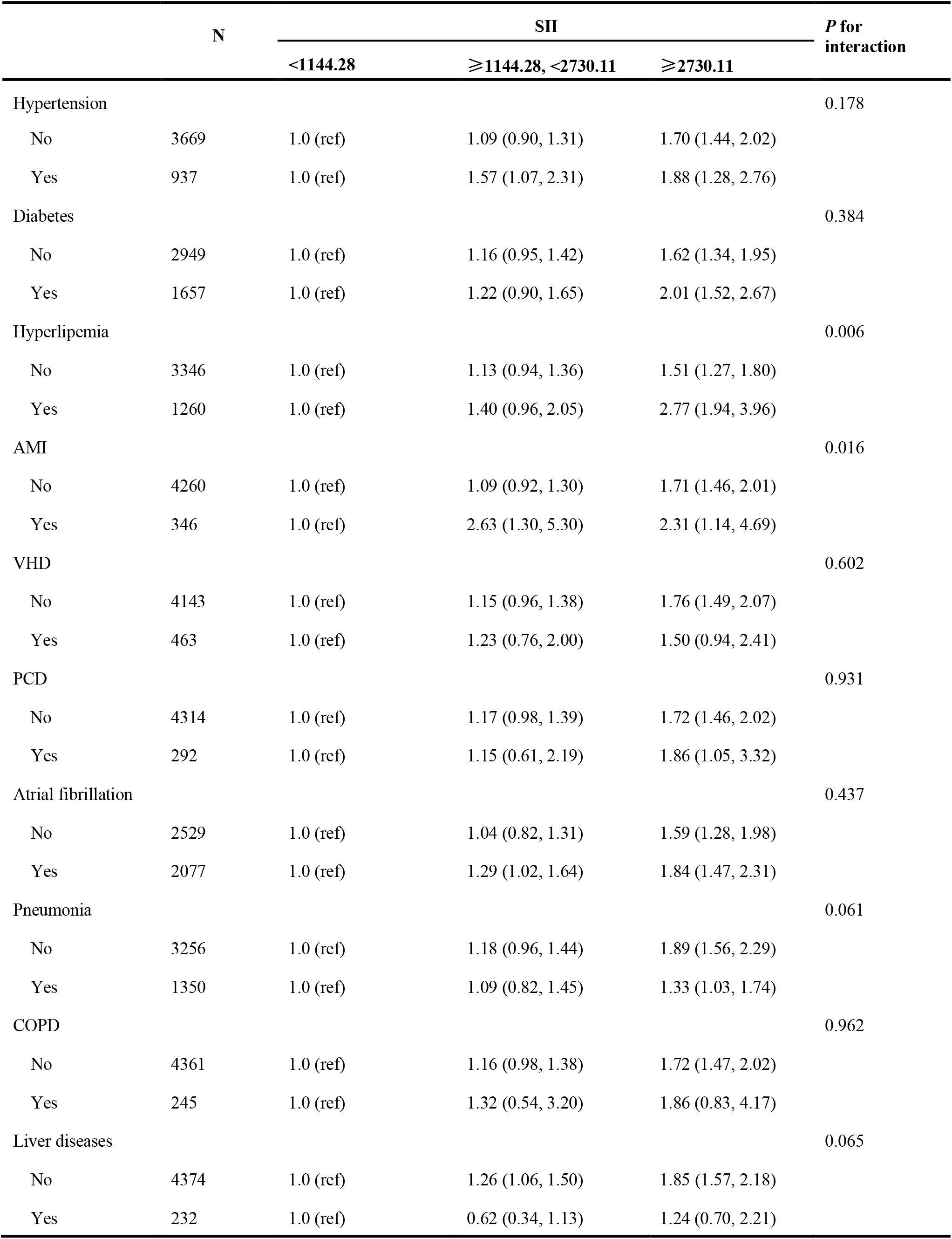
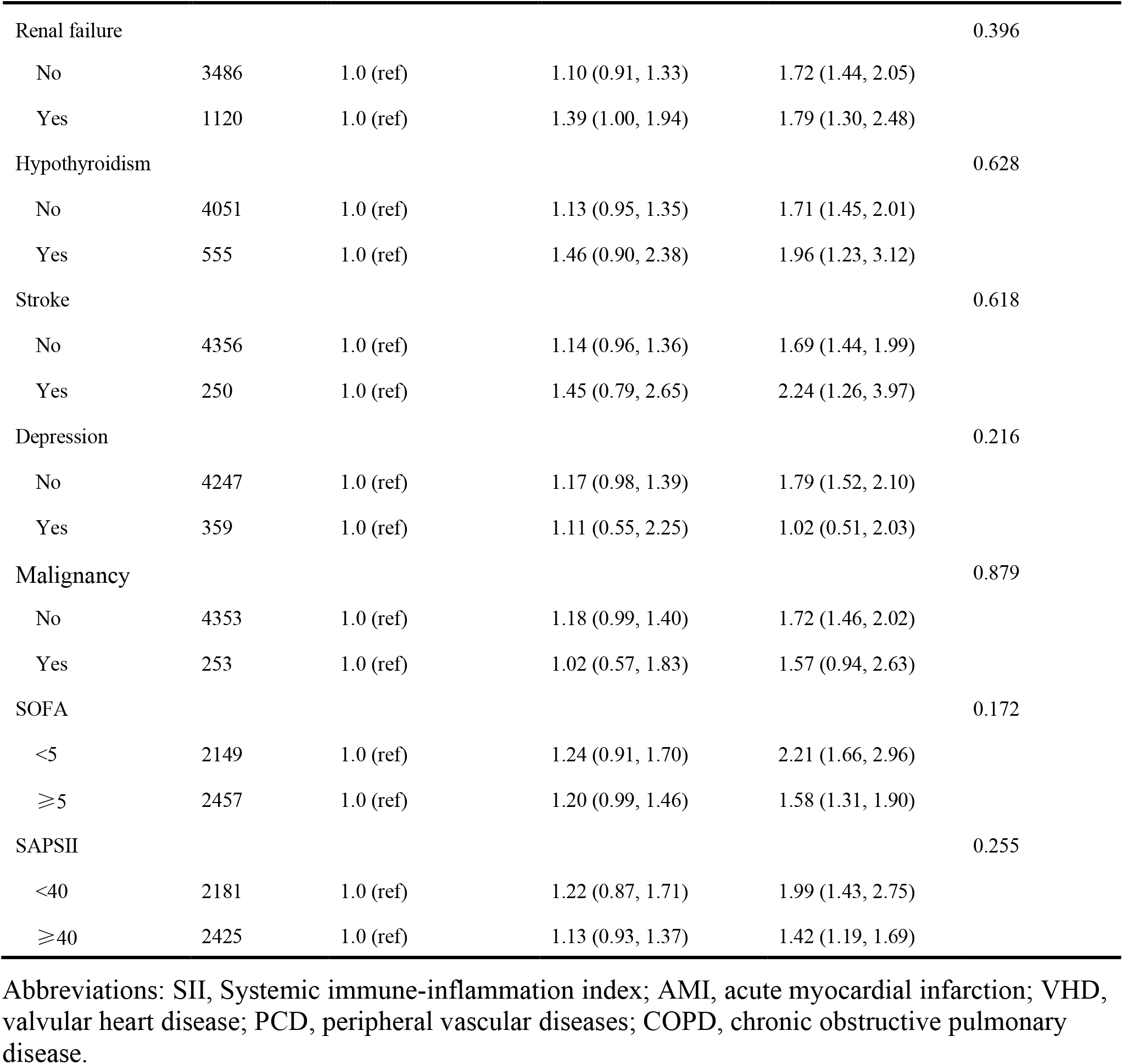
Subgroup analysis of the relationship between SII and 30-day all-cause mortality.

### 3.5 Prognostic Value of SII After PSM

Determine the optimal cut-off value of SII for 90-day all-cause mortality of critically ill patients with CHF through X-tile software, which is 5533.4. Subjects were divided into two groups according to the cut-off value, and their clinical characteristics were summarized in Table 5, most of which were unevenly distributed across the two groups. In order to effectively balance these confounding factors and improve the credibility of our results, we conducted a PSM analysis with 1:1 matching. After PSM, 602 pairs of research objects were generated and the difference of almost all variables were balanced between the two groups, with a good matching performance (Figure 3). At the same time, after PSM, compared with the low SII level group, subjects in the high SII level group still underwent higher 30- (24.9% vs 33.2%, *p*=0.002) and 90-day (35.0% vs 42.0%, *p*=0.013) and hospital (22.4% vs 27.7%, *p*=0.033) all-cause mortalities, with shorter survival time (Table 5 and Figure 4). Besides, after adjusting for these covariable in the model II, the increased SII was also associated with increased risk of short-term all-cause mortality in critically ill patients with CHF after PSM and the results were shown in Figure 5 in the form of a forest map.

**Table 5.** The clinical characteristics in critically ill patients with CHF before and after PSM.

| Parameter | Before PSM |  |  | After PSM |  |  |
| --- | --- | --- | --- | --- | --- | --- |
|  | <5533.4 | ≥5533.4 | *P* value | <5533.4 | ≥5533.4 | *P* value |
| N | 3995 | 611 |  | 602 | 602 | 602 |
| **Demographics** |  |  |  |  |  |  |
| Age, years | 74.4 (63.4-82.9) | 76.9 (68.2-83.7) | <0.001 | 77.7 (66.1-84.2) | 76.9 (68.1-83.7) | 0.984 |
| Male, n (%) | 2154 (53.9%) | 282 (46.2%) | <0.001 | 279 (46.3%) | 279 (46.3%) | 1.000 |
| Ethnicity, n (%) |  | 0.001 | 0.001 |  |  | 0.526 |
| White | 2879 (72.1%) | 470 (76.9%) |  | 463 (76.9%) | 461 (76.6%) |  |
| Black | 309 (7.7%) | 23 (3.8%) |  | 30 (5.0%) | 23 (3.8%) |  |
| Others | 807 (20.2%) | 118 (19.3%) |  | 109 (18.1%) | 118 (19.6%) |  |
| **Vital signs** |  |  |  |  |  |  |
| HR, beats/minute | 84.5 (74.1-96.0) | 88.2 (77.0-99.6) | <0.001 | 87.5 (77.3-98.8) | 87.9 (76.8-99.2) | 0.840 |
| RR, times/minute | 19.3 (16.8-22.3) | 20.3 (17.6-23.2) | <0.001 | 20.2 (17.3-23.5) | 20.2 (17.6-23.1) | 0.876 |
| SBP, mmHg | 113.8 (104.6-126.0) | 113.8 (104.6-126.0) | 0.145 | 113.9 (103.4-125.6) | 112.1 (103.5-125.7) | 0.964 |
| DBP, mmHg | 57.1 (51.1-63.9) | 55.7 (50.2-62.5) | 0.009 | 55.3 (49.1-61.9) | 55.6 (50.2-62.5) | 0.219 |
| Temperature, °C | 36.8 (36.4-37.2) | 36.7 (36.3-37.1) | 0.010 | 36.8 (36.4-37.2) | 36.7 (36.3-37.1) | 0.254 |
| SpO<sub>2</sub>, % | 97.3 (95.8-98.5) | 97.0 (95.5-98.4) | 0.157 | 97.3 (95.6-98.5) | 97.0 (95.5-98.4) | 0.474 |
| Weight, kg | 78.4 (65.0-94.3) | 74.4 (61.0-90.0) | <0.001 | 73.0 (61.0-88.5) | 74.4 (61.0-90.0) | 0.560 |
| **Therapies, n (%)** |  |  |  |  |  |  |
| ACEI | 1117 (28.0%) | 149 (24.4%) | 0.065 | 153 (25.4%) | 147 (24.4%) | 0.689 |
| ARB | 158 (4.0%) | 25 (4.1%) | 0.872 | 26 (4.3%) | 25 (4.2%) | 0.886 |
| β-blocker | 2454 (61.4%) | 373 (61.0%) | 0.858 | 373 (62.0%) | 367 (61.0%) | 0.722 |
| Digoxin | 411 (10.3%) | 64 (10.5%) | 0.888 | 85 (14.1%) | 63 (10.5%) | 0.053 |
| Furosemide | 2766 (69.2%) | 424 (69.4%) | 0.937 | 412 (68.4%) | 416 (69.1%) | 0.804 |
| Statins | 1274 (31.9%) | 177 (29.0%) | 0.148 | 158 (26.2%) | 174 (28.9%) | 0.302 |
| Dialysis | 426 (10.7%) | 52 (8.5%) | 0.104 | 86 (14.3%) | 51 (8.5%) | 0.001 |
| Vasopressor | 1042 (26.1%) | 192 (31.4%) | 0.008 | 171 (28.4%) | 187 (31.1%) | 0.313 |
| Ventilation | 1561 (39.1%) | 318 (52.0%) | <0.001 | 306 (50.8%) | 310 (51.5%) | 0.818 |
| Assisted circulation | 259 (6.5%) | 23 (3.8%) | 0.009 | 21 (3.5%) | 23 (3.8%) | 0.759 |
| **Scores** |  |  |  |  |  |  |
| SOFA | 5.0 (3.0-7.0) | 5.0 (3.0-7.0) | 0.556 | 5.0 (3.0-7.0) | 5.0 (3.0-7.0) | 0.405 |
| SAPSII | 40.0 (32.0-49.0) | 45.0 (36.0-53.5) | <0.001 | 44.0 (35.0-53.0) | 45.0 (36.0-53.0) | 0.440 |

**Table 5.** Continued.
| Parameter | Before PSM |  |  | After PSM |  |  |
| --- | --- | --- | --- | --- | --- | --- |
|  | <5533.4 | ≥5533.4 | *P* value | <5533.4 | ≥5533.4 | *P* value |
| N | 3995 | 611 |  | 602 | 602 |  |
| **Laboratory events** |  |  |  |  |  |  |
| Hemoglobin, g/dl | 11.0 (9.7-12.4) | 10.7 (9.5-12.1) | 0.008 | 10.7 (9.5-11.9) | 10.7 (9.5-12.1) | 0.290 |
| Creatinine, mg/dl | 1.2 (0.9-1.8) | 1.2 (0.9-2.1) | 0.250 | 1.3 (0.9-2.0) | 1.2 (0.9-2.1) | 0.160 |
| BUN, mg/dl | 26.0 (18.0-42.0) | 30.0 (19.0-49.0) | <0.001 | 31.0 (20.0-49.0) | 30.0 (19.0-48.0) | 0.613 |
| Glucose, mg/dl | 134.0 (114.4-163.6) | 145.5 (119.7-176.8) | <0.001 | 141.2 (117.1-180.3) | 145.5 (119.6-176.2) | 0.610 |
| Sodium, mmol/L | 139.0 (136.0-141.0) | 138.0 (135.0-141.0) | <0.001 | 138.0 (135.0-141.0) | 138.0 (135.0-141.0) | 0.487 |
| Potassium, mmol/L | 4.2 (3.8-4.6) | 4.2 (3.7-4.7) | 0.575 | 4.2 (3.8-4.7) | 4.2 (3.7-4.7) | 0.634 |
| Chloride, mmol/L | 103.0 (100.0-107.0) | 102.0 (98.0-107.0) | <0.001 | 103.0 (99.0-106.0) | 102.0 (98.0-107.0) | 0.377 |
| Bicarbonate, mmol/L | 25.0 (21.0-28.0) | 24.0 (20.0-27.0) | <0.001 | 24.0 (20.0-27.0) | 24.0 (20.0-27.0) | 0.878 |
| PT, second | 14.3 (13.2-16.8) | 14.4 (13.2-17.1) | 0.245 | 14.4 (13.2-17.3) | 14.4 (13.2-17.2) | 0.846 |
| APTT, second | 31.8 (27.3-41.1) | 31.6 (27.3-38.6) | 0.329 | 31.3 (27.3-40.2) | 31.7 (27.3-38.7) | 0.920 |
| Lactate, mmol/L | 1.9 (1.3-2.9) | 1.8 (1.3-2.8) | 0.654 | 1.8 (1.3-3.0) | 1.8 (1.3-2.8) | 0.654 |
| Albumin, g/dL | 3.0 (2.6-3.4) | 2.9 (2.5-3.3) | <0.001 | 2.8 (2.4-3.3) | 2.9 (2.5-3.3) | 0.255 |
| Bilirubin, mg/dL | 1.5 (0.6-3.1) | 0.9 (0.4-2.5) | <0.001 | 1.1 (0.5-2.4) | 0.9 (0.4-2.6) | 0.282 |
| ALT, IU/L | 62.0 (22.0-309.0) | 50.0 (21.0-242.7) | 0.034 | 51.0 (21.0-225.8) | 50.0 (21.0-248.5) | 0.637 |
| NT-proBNP, ng/ml | 10.9 (4.6-18.6) | 12.1 (5.1-19.8) | 0.019 |  |  |  |
| cTnT, ng/mL | 1.4 (0.1-2.9) | 1.1 (0.1-2.5) | 0.016 | 1.1 (0.1-2.8) | 1.1 (0.1-2.5) | 0.768 |
| CI, L/min/m<sup>2</sup> | 2.8 (2.1-3.5) | 2.8 (2.2-3.5) | 0.309 | 2.8 (2.1-3.4) | 2.8 (2.2-3.5) | 0.938 |
| Urine output, L | 1.6 (0.9-2.5) | 1.4 (0.8-2.1) | <0.001 | 1.4 (0.7-2.2) | 1.4 (0.8-2.1) | 0.522 |
| **Hospitalization type** |  |  | 0.002 |  |  | 0.238 |
| Emergency, n (%) | 3498 (87.6%) | 564 (92.3%) |  | 538 (89.4%) | 555 (92.2%) |  |
| Elective, n (%) | 338 (8.5%) | 29 (4.7%) |  | 39 (6.5%) | 29 (4.8%) |  |
| **First ICU admission** |  |  | <0.001 |  |  | 0.004 |
| CCU, n (%) | 1098 (27.5%) | 138 (22.6%) |  | 143 (23.8%) | 138 (22.9%) |  |
| **Length of ICU stay, h** | 93.0 (50.0-184.0) | 109.0 (54.0-204.0) | 0.003 | 118.5 (59.0-228.8) | 109.0 (53.0-201.8) | 0.128 |
| **Comorbidities, n (%)** |  |  |  |  |  |  |
| Hypertension | 830 (20.8%) | 107 (17.5%) | 0.062 | 126 (20.9%) | 106 (17.6%) | 0.144 |
| Hyperlipemia | 1120 (28.0%) | 140 (22.9%) | 0.008 | 133 (22.1%) | 138 (22.9%) | 0.730 |
| Diabetes mellitus | 1449 (36.3%) | 208 (34.0%) | 0.285 | 205 (34.1%) | 205 (34.1%) | 1.000 |

**Table 5.** Continued.
| Parameter | Before PSM |  |  | After PSM |  |  |
| --- | --- | --- | --- | --- | --- | --- |
|  | <5533.4 | ≥5533.4 | *P* value | <5533.4 | ≥5533.4 | *P* value |
| N | 3995 | 611 |  | 602 | 602 |  |
| **Comorbidities, n (%)** |  |  |  |  |  |  |
| Atrial fibrillation | 1783 (44.6%) | 294 (48.1%) | 0.107 | 281 (46.7%) | 290 (48.2%) | 0.603 |
| AMI | 301 (7.5%) | 45 (7.4%) | 0.882 | 39 (6.5%) | 44 (7.3%) | 0.570 |
| VHD | 402 (10.1%) | 61 (10.0%) | 0.952 | 88 (14.6%) | 61 (10.1%) | 0.018 |
| Peripheral vascular | 511 (12.8%) | 66 (10.8%) | 0.167 | 88 (14.6%) | 64 (10.6%) | 0.037 |
| Pulmonary circulation | 239 (6.0%) | 53 (8.7%) | 0.011 | 45 (7.5%) | 52 (8.6%) | 0.459 |
| Pneumonia | 1108 (27.7%) | 242 (39.6%) | <0.001 | 235 (39.0%) | 237 (39.4%) | 0.906 |
| COPD | 188 (4.7%) | 57 (9.3%) | <0.001 | 48 (8.0%) | 53 (8.8%) | 0.603 |
| Liver diseases | 214 (5.4%) | 18 (2.9%) | 0.011 | 20 (3.3%) | 18 (3.0%) | 0.742 |
| Renal failure | 982 (24.6%) | 138 (22.6%) | 0.284 | 162 (26.9%) | 136 (22.6%) | 0.083 |
| Stroke | 227 (5.7%) | 23 (3.8%) | 0.051 | 38 (6.3%) | 23 (3.8%) | 0.052 |
| Hypothyroidism | 461 (11.5%) | 94 (15.4%) | 0.007 | 97 (16.1%) | 92 (15.3%) | 0.692 |
| Malignancy | 209 (5.2%) | 44 (7.2%) | 0.047 | 31 (5.1%) | 43 (7.1%) | 0.150 |
| Depression | 303 (7.6%) | 56 (9.2%) | 0.175 | 50 (8.3%) | 56 (9.3%) | 0.542 |
| **30-day mortality** | 769 (19.2%) | 204 (33.4%) | <0.001 | 150 (24.9%) | 200 (33.2%) | 0.002 |
| **90-day mortality** | 1120 (28.0%) | 260 (42.6%) | <0.001 | 211 (35.0%) | 253 (42.0%) | 0.013 |
| **Hospital-day mortality** | 659 (16.5%) | 172 (28.2%) | <0.001 | 135 (22.4%) | 167 (27.7%) | 0.033 |

Abbreviations: PSM, propensity score matching; HR, heart rate; RR, respiratory rate; SBP, systolic blood pressure; DBP, diastolic blood pressure; SpO<sub>2</sub>, percutaneous oxygen saturation; ACEI, angiotensin-converting enzyme inhibitors; ARB, angiotensin receptor blockers; BUN, blood urea nitrogen; PT, prothrombin time; APTT, activated partial thromboplastin time; ALT, alanine transaminase; cTnT, troponin T; NT-proBNP, N-terminal pro brain natriuretic peptide; CI, cardiac index; SII, Systemic immune-inflammation index; CHF, congestive heart failure; AMI, acute myocardial infarction; VHD, valvular heart disease; COPD, chronic obstructive pulmonary disease. CCU, cardiac care unit.

**Figure 3.**
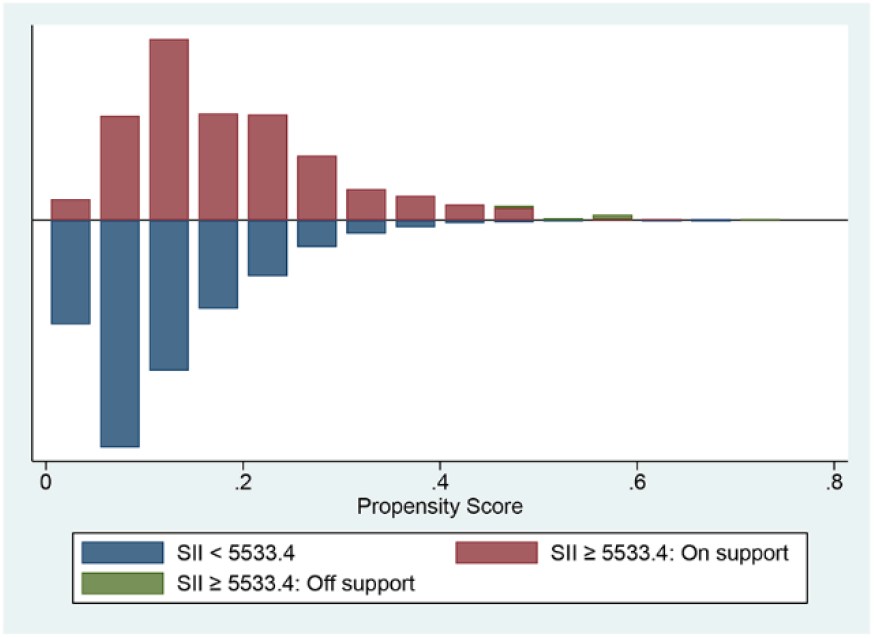
The histogram presenting the range of propensity scores and the corresponding number of matches in high (red) and low (blue) SII groups. SII, systemic immune-inflammation index.

**Figure 4.**
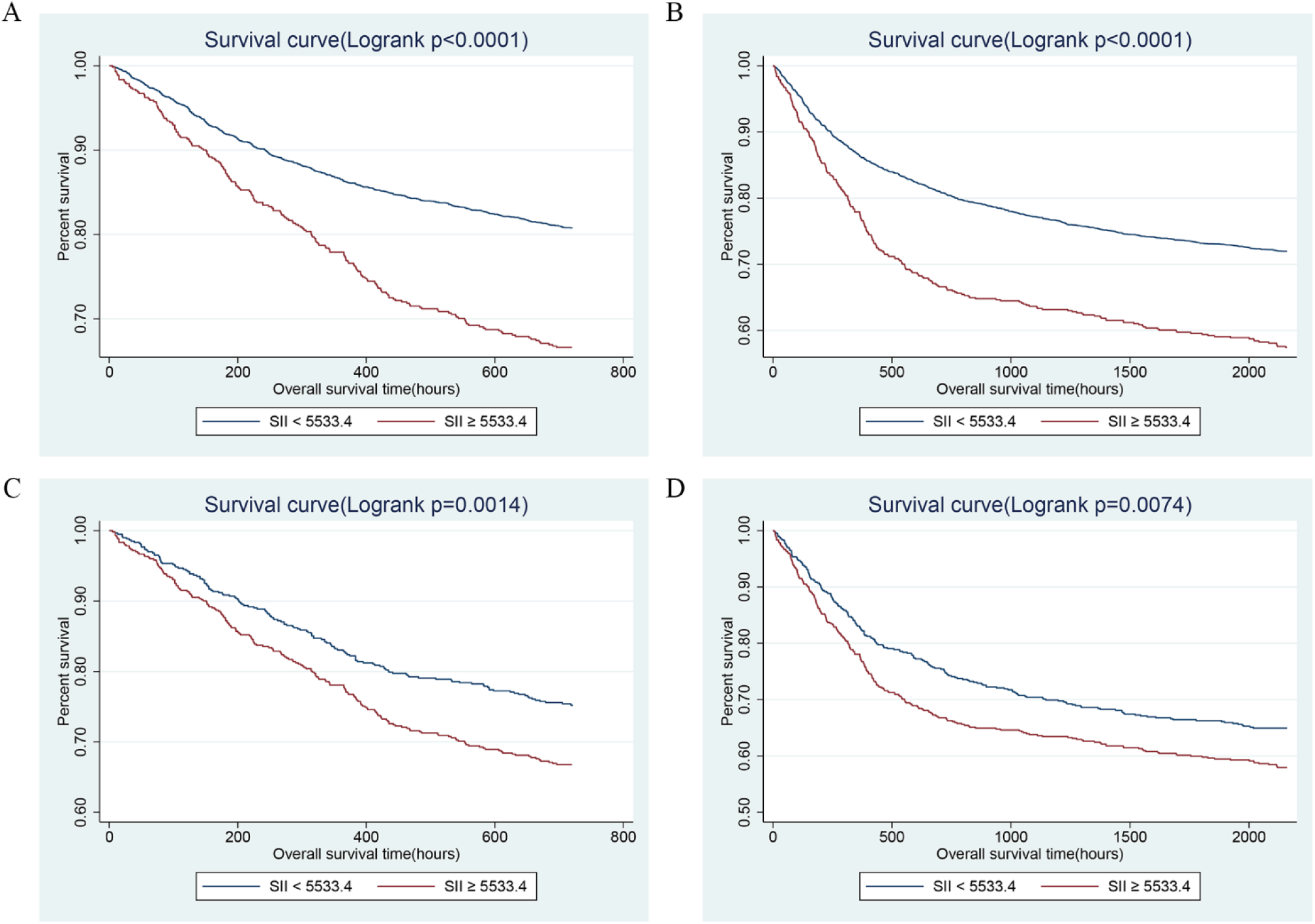
Kaplan-Meier survival curves showing the 30- (A) and 90- (B) day all-cause mortalities stratified by cut-off value of SII in critically ill patients with CHF before PSM. C and D presented the correlation between SII and 30- and 90-day mortalities after PSM, separately. SII, systemic immune-inflammation index. CHF, congestive heart failure. PSM, propensity score matching.

**Figure 5.**
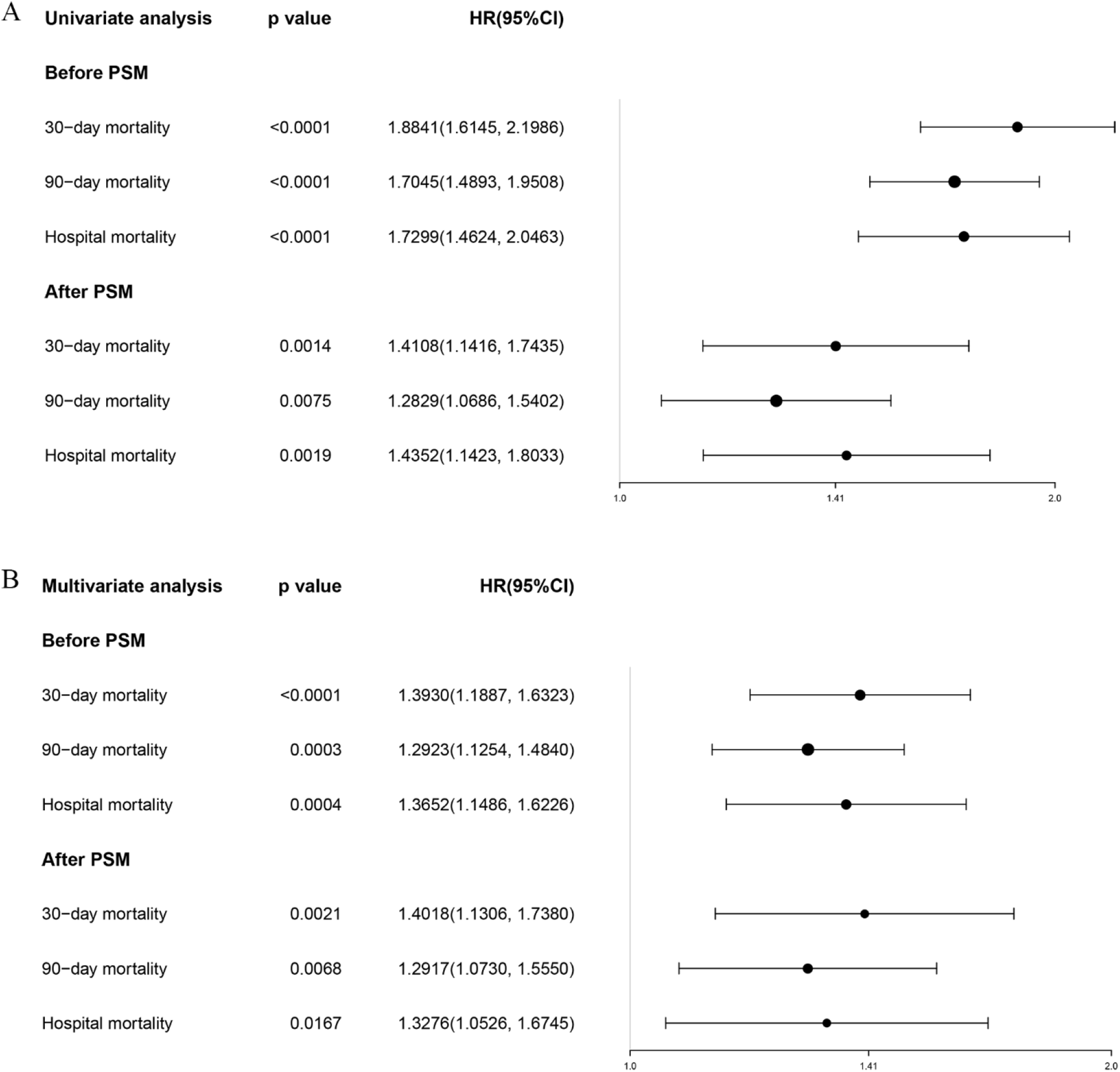
The forest plot showing the univariate (A) and multivariate (B) Cox regression for effects of SII on short-term mortality in the critically ill patients with CHF before and after PSM. SII, systemic immune-inflammation index. CHF, congestive heart failure. PSM, propensity score matching.

## 4 Discussion

CHF is widespread all over the world, with gradually increasing incidence with age, and has become one of the diseases with the highest rates of hospitalizations and deaths in the world, bringing about enormous medical expenditure and social burden, especially in developing countries(Savarese and Lund, 2017). In addition to exploring promising treatments, the development of novel prognostic markers for risk stratification of patients is also of great value for improving the prognosis of patients. As we all know, as a commonly used indicator of heart failure, NT-proBNP can make a good performance in prognostic evaluation. However, the clinical applications of NT-proBNP for risk assessment has some drawbacks and problems. First, compared to the real value, the detection of NT-proBNP tend to be underestimated because of the short half-life. Moreover, the test of NT-proBNP may be affected by various factors such as age, gender, and kits(Cho et al., 2020). Therefore, the search for more possible biomarkers has gradually attracted the attention of researchers and clinicians.

In the present study, we extracted the clinical data of 4606 critically ill patients who diagnosed with CHF between 2001 and 2012 in MIMIC-III database, and analyzed the relationship between SII and short-term prognosis by univariate and multivariate regression analyses for the first time. And the results have verified that SII could be an independent biomarker for poor short-term prognosis of CHF. There was a nonlinear relationship between SII and 30- and 90-day mortalities. And the higher SII level, the higher risk of 30- and 90-day and hospital mortalities, as well as the higher incidence of MACEs. The correlation remained significant after adjusting for possible confounders, stratifying according to comorbidities, and PSM matching, respectively.

Inflammation, a widely accepted hallmark of heart failure, has been proven to play an indispensable role in the pathogenesis of heart failure(Dick and Epelman, 2016). The increase and activation of inflammation-related cytokines not only present a reflection of the activation of inflammation in vivo, but have also been identified as being associated with poor prognosis in heart failure. Tumor necrosis factor α (TNF-α) was the first cytokine found to be significantly elevated in the serum of patients with heart failure, which could directly induce myocardial apoptosis and necrosis, bringing about ventricular adverse remodeling(Anker and von Haehling, 2004). The elevated levels of TNF-α often indicated impaired cardiac systolic function and poor long-term survival(Rauchhaus et al., 2000). In addition, interleukin 6 (IL-6), another classic cytokine mainly derived from monocytes, could be significantly increased in patients with left ventricular systolic dysfunction without clinical symptoms, and may be a sensitive indicator for the early diagnosis of heart failure(Plenz et al., 2001). IL-6 was also positively correlated with the severity of heart failure, the concentration of which was negatively correlated with left ventricular ejection fraction and overall survival(Markousis-Mavrogenis et al., 2019). Although these inflammatory molecules had good performance in the prognostic evaluation of patients, the expensive detection cost limited their clinical application. As a novel biomarker, SII can systematically and comprehensively reflect the status of inflammation in vivo. Blood routine is a routine examination for almost all hospitalized patients. It is simple, fast, and inexpensive. Compared with NT-proBNP, cytokines and other prognostic indicators, SII can effectively screen high-risk patients, guide the formulation of individualized treatment plans and improve the prognosis of patients without additional cost, with a broad application prospect, especially in underdeveloped areas.

The mechanisms of the relationship between SII and adverse prognosis of CHF remains unclear, and possible explanations are as follows. Recently, it has been revealed that circulating immune cells and their subtypes had a vital indicative effect on the prognosis of cardiovascular diseases(Strassheim et al., 2019). SII was a prognostic indicator that integrates three circulating immune cells, including neutrophils, lymphocytes, and platelets, the increase of which indicates either a relative increase in platelets and neutrophils counts or a relative decrease in lymphocytes. Neutrophils are the main component of white blood cells, accounting for 60-70% of the total, and play an important role in the non-specific immune system. In the inflammatory response, neutrophils react rapidly, with strong chemotaxis and phagocytosis, and participate in the progression of various cardiovascular diseases, including heart failure(Bonaventura et al., 2019). In addition, neutrophils could contribute to oxidative stress and endothelial dysfunction by releasing a large amount of myeloperoxidase, NADPH oxidase, and so on(Swirski and Nahrendorf, 2013). According to a community-based study(Arruda-Olson et al., 2009), absolute neutrophils can effectively predict the increased incidence of acute decompensated heart failure after AMI, and it was also an independent risk factor for death(Uthamalingam et al., 2011). Lymphocytes mainly consist of two subtype including T and B cells, which were often associated with the specific immunity. Several previous studies have reported that lymphocyte counts in patients with heart failure were lower than in the normal population and lymphopenia served as an independent predictor of poor survival of patients with chronic and advanced heart failure(Charach et al., 2011; Vaduganathan et al., 2012). Lymphopenia was considered to be an important feature of systemic inflammation, which may be caused by the following mechanisms. First, the circulating lymphocytes were attracted into the cardiac tissue, leading to its redistribution. Then, lymphopenia was related to the activation of renin-angiotensin-aldosterone hormone and the adrenergic nervous system, which could exert pro-apoptotic effects on lymphocytes(Charach et al., 2011). Platelets were differentiated from the mononuclear-phagocyte system and were the key mediator linking the two pathological processes of inflammation and thrombosis(Ye et al., 2019). On the one hand, platelets were the main effector molecules for hemostasis after cardiovascular injury. On the other hand, the activation of platelets plays a major role in pathogenic thrombosis and participates in the pathogenesis of many groups of cardiovascular diseases, including coronary heart disease(Glezeva et al., 2016). Moreover, Platelets interacted with leukocytes and their subtypes (neutrophil, lymphocytes, etc.) and endothelial cells and activated them, inducing monocyte adhesion and transport, releasing inflammatory factors such as TNF-α and IL-1, which together promoted local inflammation and fibrosis in heart failure(Glezeva et al., 2016). The study by Kandis(Kandis et al., 2011) et al. demonstrated that mean platelet volume (MPV), indicating the activation of platelets, increased significantly in decompensated heart failure patients. In addition, MPV at admission was independently associated with the hospital and 6-month mortalities.

Besides, the subgroup analysis was also conducted, and the results demonstrated that the association was stable and consistent between the high level of SII and the poor 30-day mortality across CHF patients with different comorbidities or severity scores. We also noted that, among the critically ill patients with CHF, patients complicated with AMI and hyperlipidemia had a higher risk of 30-day mortality, and this risk was higher for higher SII, which implied that SII may be more valuable for the prognostic evaluation of CHF patients with AMI or hyperlipidemia. Similarly, the previous study has also shown that in patients with coronary heart disease, the high level of SII indicated a high risk of cardiac death in the future, as well as the high incidence of non-fatal stroke and heart failure(Yang et al., 2020). Although there was no research on the correlation between SII and hyperlipidemia, the interconnection of hyperlipidemia and inflammation involved in the pathogenesis of cardiovascular disease has been recognized(Rodriguez-Garcia and Alcaide, 2021).

As a retrospective cohort study, the existence of confounding factors cannot be ignored, and it was difficult to intuitively and accurately judge the correlation between SII level and the prognosis of CHF patients. Another highlight of this study was the use of PSM analysis to reduce the imbalance of confounding factors across different groups. After PSM, the most baseline characteristics were well-balanced between these two groups, except for the use of dialysis, the type of first ICU admission, and the peripheral vascular diseases. More importantly, the conclusion that high levels of SII were independently related to high short-term mortality remained stand before and after PSM, which improved the reliability of SII as a prognostic marker of CHF.

But at the same time, there were certain limitations in this study. First, the present study was conducted at a single center and did not validate the prognostic value of SII in a validation cohort. Second, the clinical data were collected retrospectively from databases and it was difficult to ensure that variables were evenly distributed across groups, although multiple regression models were conducted to adjust for confounders and PSM analysis was used to minimize inter-group differences in baseline characteristics. Third, some significant variables may be omissive due to the lack of data. Last, this study was unable to determine the underlying mechanism of the association between high SII and poor prognosis of CHF patients, and further experiments are necessary.

## 5 Conclusion

In summary, the high level of SII is closely related to the poor short-term prognosis in critically ill patients with CHF, including 30- and 90-day and hospital all-cause mortalities, as well as the occurrence of MACEs, and is expected to be a simple and effective prognostic evaluation indicator.

## Data Availability

Publicly available datasets were analyzed in this study. This data can be extracted from Monitoring in Intensive Care Database III version 1.4 (MIMIC-III v.1.4) after passing on the required courses and obtaining the authorization.

https://mimic.mit.edu/

## 7 Author Contributions

Zaixin Yu and Lihuang Zha are the senior author who conceived and designed the study. Yiyang Tang and Xiaofang Zeng collected and analyzed the clinical data. Yiyang Tang drafted the manuscript. Yilu Feng and Qin Chen participated in the implementation of statistical methods in this study and put forward constructive suggestions. Zhenghui Liu and Hui Luo reviewed the study and participated in the interpretation of the results. All authors gave final approval of the version to be published, and agree to be accountable for all aspects of the work.

